# Drug utilization pattern and cost analysis of anticancer drugs among cancer patients attending a tertiary cancer hospital in Nepal: A cross-sectional study

**DOI:** 10.64898/2026.07.29.26359297

**Authors:** Neha Sigdel, Anil Bhusal, Krishna Neupane, Prabha Adhikari, Radhika Thapa, Poonam Pant

## Abstract

Cancer imposes a rising and largely out-of-pocket-financed burden in Nepal, yet patient-level data on how anticancer drugs are prescribed and what chemotherapy actually costs patients remain limited. This study described the drug utilization pattern and cost of anticancer drug therapy among cancer patients treated at a tertiary cancer hospital in Nepal.

A descriptive, cross-sectional, record-based study was conducted at Bhaktapur Cancer Hospital, Bhaktapur, Nepal. Medical records of cancer patients aged above 18 years attending the day-care chemotherapy ward were reviewed using a structured proforma. Sociodemographic characteristics, diagnosis, regimen and cycle data, and WHO/INRUD prescribing indicators were recorded, along with the actual cost, maximum retail price, and Nepal Health Insurance Board reimbursement rate for anticancer drug therapy. Data were analyzed descriptively and, given the skewed distribution of cost, on a log-transformed cost variable using Pearson correlation, independent-samples t tests, and multiple linear regression in SPSS version 16.

A total of 151 chemotherapy patients were analyzed (mean age 53.81 ± 12.51 years; 67.5% female). Breast cancer (26.5%) was the most common diagnosis. Patients received a mean of 10.56 ± 2.19 total drugs, of which 1.72 ± 0.67 were anticancer agents; 99.85% of drugs were prescribed generically, and 63.2–67.7% were on the national essential medicines list. Mean actual cost of anticancer therapy per patient was NPR 15,245.28 ± 15,672.33, substantially below the mean maximum retail price (NPR 38,803.22). In bivariate analysis, log-transformed cost was significantly higher among male patients (p =.038) and among patients not fully compliant with the essential medicines list (p =.028), but no patient, disease, or prescribing variable — including sex — independently predicted log-transformed cost in a multiple regression model that was not statistically significant overall (R² =.113, p =.323).

Anticancer prescribing at this hospital was characterized by near-universal generic use but only partial alignment with the national essential medicines list, and chemotherapy cost varied widely across patients without being reliably explained by the patient or prescribing factors examined. These findings support continued WHO/INRUD-style prescribing surveillance and closer attention to essential-medicines-list adherence and drug pricing in this setting.

## Introduction

Cancer is an escalating public health concern in Nepal: GLOBOCAN 2020 estimated 20,508 new cancer cases and 13,629 cancer-related deaths nationally, with an age-standardized incidence rate of 80.9 and mortality rate of 54.8 per 100,000 population[1]. Chemotherapy remains a cornerstone of treatment for most solid and hematologic malignancies, typically combining one or more anticancer agents with adjuvant and supportive drugs across multiple cycles. Because these regimens are complex, costly, and carry a narrow therapeutic index, periodic evaluation of how anticancer drugs are actually prescribed — and what that prescribing costs patients — is an important complement to clinical guideline development.

Consistent with global trends, in which cancer accounted for an estimated 9.6 million deaths in 2019 and the burden continues to concentrate in low- and middle-income countries where health systems are least equipped to absorb it[2], Nepal’s cancer burden is shifting toward the resource-intensive, chronic-management model typical of non-communicable disease care, in which repeated, multi-drug chemotherapy cycles rather than single curative interventions define much of clinical practice.

Chemotherapy regimens for common malignancies increasingly combine multiple cytotoxic, targeted, and supportive agents across several cycles, and this complexity raises the practical importance of monitoring not only clinical outcomes but also how these drugs are actually selected, combined, and paid for at the point of care.

The World Health Organization/International Network for Rational Use of Drugs (WHO/INRUD) core prescribing indicators — average drugs per encounter, percentage prescribed by generic name, and percentage prescribed from an essential medicines list, among others — provide a standardized framework for describing prescribing patterns and flagging potentially irrational drug use[3]. In Nepal, the National List of Essential Medicines (NLEM), most recently revised in 2021, includes a dedicated section on antineoplastic and immunomodulating agents and is the relevant benchmark against which anticancer prescribing can be assessed[4]. Hospital-based studies applying these indicators to oncology settings in Nepal remain few. A recent study at a 110-bed national referral-level cancer hospital found a mean of 8.44 ± 3.27 adverse drug reactions per enrolled patient, underscoring both the intensity of drug exposure in this population and the value of systematic, encounter-level prescribing data[5]; a separate drug utilization study of lung cancer patients at the same hospital reported an average of 9.28 drugs prescribed per encounter[6].

Since their original WHO formulation, these core prescribing indicators have been applied and cross-validated across a wide range of settings and specialties beyond oncology — including public hospitals in eastern Ethiopia[7], primary healthcare centers and tertiary hospitals in Bahawalpur, Pakistan[8], a university referral hospital in southern Ethiopia[9], and a tertiary hospital in southeastern Nigeria [10] — which consistently show that average drugs per encounter, generic-prescribing rates, and essential-medicines-list adherence vary widely by country, facility level, and case mix, underscoring the value of setting- and specialty-specific data rather than relying on indicators derived elsewhere.

Within oncology specifically, WHO/INRUD-based drug utilization evaluations have been conducted at tertiary cancer centers in India, reporting an average of 1.72–2.11 anticancer drugs and 8.77–12.22 total drugs per encounter alongside predominantly platinum- and antimetabolite-based regimens [11–13], and adverse-drug-reaction surveillance studies in Eastern Indian teaching hospitals have documented substantial toxicity burden associated with these same drug classes [14–16], reinforcing that utilization pattern and safety monitoring are complementary lenses on the same underlying prescribing practice. In Nepal, a pharmacovigilance center specifically for chemotherapy-related reactions was established at a national cancer hospital to address this gap[17], and a validated tool for predicting chemotherapy-induced neutropenia risk has since been piloted in the same setting[18], reflecting a broader institutional shift toward more systematic monitoring of anticancer drug use and its consequences.

The financial dimension of cancer treatment in Nepal is similarly under-documented at the prescribing level. Nepal’s health financing relies heavily on out-of-pocket payment, and a cross-sectional study at a tertiary cancer hospital estimated a mean direct cost of cancer treatment of NPR 387,500 per patient, with medical costs accounting for over 80% of that total and the great majority of patients reporting financial hardship as a result[19]. Compounding this burden, an assessment of anticancer medicine pricing across Nepalese hospital pharmacies found considerable brand-level price variation — in some cases exceeding 300% for the same molecule — despite government price regulation of selected products[20]. Together, these findings suggest that the cost actually paid for a course of anticancer therapy may diverge substantially from list or maximum retail prices, and that this divergence is worth quantifying alongside prescribing pattern.

Selection of which cancer medicines a country’s essential medicines list should include is itself an active area of international policy attention: the WHO Cancer Medicines Working Group has progressively tightened the evidentiary bar — including a minimum overall-survival-benefit threshold — for adding new cancer medicines to its Model List [21], a shift reflected in 2021 when WHO announced it was prioritizing access to cancer and diabetes treatments in its updated Essential Medicines Lists; yet a survey of oncologists across 82 countries found that even medicines they consider essential are inconsistently available, particularly in low- and lower-middle-income settings[22], and a related analysis found that fewer than half of the cancer medicines recommended by international treatment guidelines are typically reflected in the national essential medicines lists of low- and middle-income countries[23]. Older essential-medicines-programme evaluations reached similar conclusions decades earlier, finding that structured essential-drug programmes improve both availability and rational use where implemented consistently[24], and a widely cited set of policy recommendations for improving medicine use in developing countries specifically highlighted prescribing audits and essential-list adherence monitoring — exactly the kind of patient-level indicator collected in this study — as practical levers for improvement[25]. Nepal’s own essential medicines list has followed this international trajectory, with its most recent (sixth) revision retaining a dedicated antineoplastic and immunomodulating-agent section aligned to the WHO Model List framework.

Despite this evidence, few Nepalese studies have jointly examined anticancer drug utilization and the cost of therapy at the level of the individual chemotherapy patient, across a broad mix of cancer diagnoses rather than a single tumor type. This study therefore aimed to describe the sociodemographic and disease profile, drug prescribing pattern, and cost of anticancer drug therapy among cancer patients attending a tertiary cancer hospital in Nepal, and to examine which patient, disease, and prescribing characteristics were associated with that cost.

## Methods

### Study design and setting

This was a descriptive, cross-sectional study using quantitative methods, conducted at Bhaktapur Cancer Hospital, Dudhpati, Bhaktapur, Nepal — a well-recognized national-level cancer hospital that draws patients from across the country[5]. The study was conducted over a two-month period, from May to June 2022.

### Participants

The study population comprised patients visiting Bhaktapur Cancer Hospital for chemotherapy; the study unit was the individual patient diagnosed with cancer. Patients above 18 years of age were eligible for inclusion; patients unwilling to participate were excluded. The minimum required sample size was estimated using Cochran’s formula for an infinite population proportion:

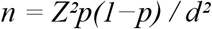

where Z is the standard normal deviate corresponding to the desired confidence level (Z = 1.96 for 95% confidence), p is the assumed prevalence of the outcome of interest (p = 9.588%, based on the brand-level price-variation proportion reported for anticancer medicines in Nepalese hospital pharmacies[20], used here in the absence of a study-specific prior estimate of prescribing or cost outcomes), and d is the desired absolute precision (d = 0.05). Substituting these values, n = (1.96)² × 0.09588 × (1 − 0.09588) / (0.05)² = 3.8416 × 0.0867 / 0.0025 ≈ 134, giving a calculated minimum target of 134 chemotherapy patients. Anticipating a non-response rate of 15%, a conservative allowance commonly applied in sample-size planning for survey- and record-based health research, this minimum target was adjusted to n / (1 − 0.15) = 134 / 0.85 ≈ 158 chemotherapy patients. The achieved sample of 151 patients therefore corresponds to a response rate of 151/158 ≈ 95.6% relative to this adjusted target.

### Data collection

Data were abstracted from patients’ medical records and prescriptions using a structured data collection form (proforma) developed for this study, after a pretest on 10% of the anticipated sample to check clarity and completeness. For each patient, the form captured sociodemographic characteristics (age, sex, ethnicity, marital status), diagnosis, chemotherapy cycle number and regimen, the anticancer and supportive drugs prescribed, and the cost of anticancer drug therapy. Printed data collection forms were used in the field, and data were subsequently entered into SPSS version 16 for analysis; all collected forms were reviewed daily for completeness and accuracy.

### Measures

Drug prescribing pattern was characterized using WHO/INRUD-adapted prescribing indicators, including the number of anticancer, adjuvant, and supportive drugs prescribed per patient; the route of administration; the percentage of drugs prescribed by generic name; and the percentage of drugs appearing on the Nepal National List of Essential Medicines (NLEM) 2021, evaluated under both a strict and a borderline-inclusive matching criterion[4]. Regimen complexity was classified as monotherapy or combination therapy, and patient-level indicators were recorded for antibiotic co-prescription, use of an additional (non-chemotherapy) injection, and use of a cytoprotective drug. Cost of anticancer drug therapy was recorded per patient as the total actual cost paid (NPR), the total maximum retail price (MRP), and the applicable Nepal Health Insurance Board (HIB) reimbursement rate.

### Statistical analysis

Data were analyzed in SPSS version 16. Descriptive statistics (means, standard deviations, minimum/maximum, frequencies, and percentages) summarized demographic, disease, and prescribing variables (N = 151 for all variables, with no missing data on any core variable). Distributional shape was assessed using skewness and kurtosis and formally tested with the Shapiro–Wilk and Kolmogorov–Smirnov tests via the EXAMINE procedure; because Total_Cost was markedly right-skewed and leptokurtic, a natural log transformation (LogCost) was applied and retained as the dependent variable for all inferential analyses of cost.

Bivariate associations between LogCost and continuous predictors (patient age, chemotherapy cycle number, total anticancer and total prescribed drugs per patient, and percentage of drugs generic or NLEM-listed) were examined with Pearson product-moment correlation. Comparisons of mean LogCost across binary prescribing and demographic indicators (sex, antibiotic co-prescription, additional injection use, cytoprotective-drug use, regimen type, all-drugs-generic status, and strict NLEM compliance) were conducted with independent-samples t tests, with Levene’s test used to check the equality-of-variances assumption for each comparison. One-way analyses of variance (ANOVA), with Tukey and Games-Howell post-hoc comparisons, were specified to compare LogCost across diagnosis, organ system, broad age band, ethnicity, and chemotherapy cycle-count group.

A standard multiple linear regression was used to examine the joint contribution of patient, disease, and prescribing characteristics — age, sex, cycle number, regimen complexity, total anticancer drugs, total drugs prescribed, antibiotic co-prescription, additional injection use, cytoprotective-drug use, percentage of drugs on the strict NLEM list, and organ-system dummy variables (reference: breast) — to LogCost. Model diagnostics included the overall model F test and R²/adjusted R², tolerance and variance inflation factors (VIF) to assess multicollinearity, the Durbin–Watson statistic to assess residual autocorrelation, and standardized residuals and Cook’s distance to identify influential cases; the normal P-P plot and histogram of standardized residuals were inspected to check the normality assumption. A significance threshold of α =.05 (two-tailed) was used throughout.

### Ethical consideration

Ethical approval was obtained from the Institutional Review Committee (IRC) of CIST College, Pokhara University (Approval No. IRC/114/078/79), and administrative approval was subsequently obtained from Bhaktapur Cancer Hospital. Because data were abstracted from existing medical records, participants were informed of the study objective and both written and verbal consent were obtained from patients where applicable. Confidentiality of participant information was maintained throughout data collection, analysis, and reporting, and data were used solely for research purposes. Participation was voluntary throughout.

## Results

### Drug utilization pattern

#### Sample Characteristics

A total of 151 chemotherapy patients were included in the analysis, with no missing data on any of the core demographic or clinical variables. Patients ranged in age from 19 to 78 years (M = 53.81, SD = 12.51). The majority of the sample was female (67.5%, n = 102), with males comprising 32.5% (n = 49). The most represented age group was 40–59 years (51.0%), followed by ≥60 years (34.4%); patients aged 18–39 years were least represented (14.6%). By ethnic/social group, the sample was composed of Janajati (38.4%), Brahmin/Chhetri (35.8%), Newar (22.5%), and a small combined ‘Others’ category comprising Dalit, Madhesi, and unknown classifications (3.3%). Breast cancer was the most common diagnosis (26.5%), followed by lung cancer (16.6%) and ovarian cancer (10.5%); no other single diagnosis exceeded 7% of the sample, and diagnoses with fewer than five cases were collapsed into an ‘Others’ category (21.9%). When regrouped by organ system, breast cancer again predominated (26.5%), followed by the gynecological/reproductive system (22.5%), the gastrointestinal system (17.2%), and the respiratory system (16.6%); genitourinary and hematologic diagnoses were combined into a residual ‘Others’ category (4.6%) owing to small cell sizes. Chemotherapy cycle number ranged from 1 to 14 (M = 3.49, SD = 2.17), with cycle 1 (20.5%) and cycle 3 (20.5%) the most frequently recorded and cycles of 7 or more collapsed into a single category (8.7%). Regimen complexity was classified as combination therapy for 60.3% of patients (n = 91) and monotherapy for 39.7% (n = 60); no patients had regimen type unrecorded. The demographic and clinical characteristics of the study participants are presented in Table 1.

**Table 1.** Frequency Distribution of Demographic and Disease-Related Variables (N = 151)

| Variable / Category | n | % |
| --- | --- | --- |
| **Sex** |  |  |
| Male | 49 | 32.5 |
| Female | 102 | 67.5 |
| **Age Group** |  |  |
| 18–39 | 22 | 14.6 |
| 40–59 | 77 | 51.0 |
| ≥60 | 52 | 34.4 |
| **Ethnicity / social group** |  |  |
| Brahmin/Chhetri | 54 | 35.8 |
| Janajati | 58 | 38.4 |
| Newar | 34 | 22.5 |
| Others (Dalit/Madhesi/Unknown) | 5 | 3.3 |
| **Cancer diagnosis** |  |  |
| Breast | 40 | 26.5 |
| Cervical | 10 | 6.6 |
| Esophagus | 5 | 3.3 |
| Lung | 25 | 16.6 |
| Nasopharynx | 6 | 4.0 |
| Ovarian | 16 | 10.5 |
| Rectum | 5 | 3.3 |
| Stomach | 6 | 4.0 |
| Supraglottis | 5 | 3.3 |
| Others (<5 cases each) | 33 | 21.9 |
| **Organ system group** |  |  |
| Breast | 40 | 26.5 |
| Gastrointestinal | 26 | 17.2 |
| Gynecological (Reproductive) | 34 | 22.5 |
| Head & Neck | 19 | 12.6 |
| Respiratory | 25 | 16.6 |
| Others (Genitourinary/Hematologic) | 7 | 4.6 |
| **Chemotherapy cycle group** |  |  |
| 1 | 31 | 20.5 |
| 2 | 23 | 15.2 |
| 3 | 31 | 20.5 |
| 4 | 27 | 17.9 |
| 5 | 14 | 9.3 |
| 6 | 12 | 7.9 |
| $\geq 7$ | 13 | 8.7 |
| **Regimen complexity** |  |  |
| Monotherapy | 60 | 39.7 |
| Combination therapy | 91 | 60.3 |

*Note. N = 151 for all variables with no missing data. Percentages are column (valid) percentages.*

**Table 2.** Descriptive Statistics for Continuous Prescribing Indicators (N = 151)

| Variable | Min | Max | M | SD |
| --- | --- | --- | --- | --- |
| Total anticancer drugs / patient | 1.00 | 4.00 | 1.72 | 0.67 |
| IV anticancer drugs / patient | 1.00 | 4.00 | 1.60 | 0.68 |
| Oral anticancer drugs / patient | 0.00 | 1.00 | 0.11 | 0.32 |
| Adjuvant (injectable) drugs / patient | 0.00 | 7.00 | 3.70 | 1.15 |
| Oral supportive drugs / patient | 0.00 | 8.00 | 5.15 | 1.45 |
| Total drugs / patient (all categories) | 3.00 | 16.00 | 10.56 | 2.19 |
| % drugs generic / patient | 91.67 | 100.00 | 99.85 | 1.08 |
| % drugs on NLEM<br>2021, strict | 25.00 | 100.00 | 63.17 | 20.28 |
| % drugs on NLEM<br>2021, incl.<br>borderline | 33.33 | 100.00 | 67.67 | 17.56 |

**Table 3.** Frequency of Patient-Level Drug-Related Prescribing Indicators (N = 151)

| Indicator | No / 0<br>(n) | % | Yes / 1<br>(n) | % |
| --- | --- | --- | --- | --- |
| Antibiotic co-prescribed | 148 | 98.0 | 3 | 2.0 |
| Additional injection used | 141 | 93.4 | 10 | 6.6 |
| Cytoprotective drug used | 110 | 72.8 | 41 | 27.2 |
| All drugs generic (patient-level) | 3 | 2.0 | 148 | 98.0 |
| All drugs on NLEM 2021, strict (patient-level) | 146 | 96.7 | 5 | 3.3 |

*Note. “All drugs generic” and “All drugs on NLEM, strict” are patient-level composite indicators, coded 1 only when* *every drug for that patient met the respective criterion.*

**Table 4.** WHO/INRUD-Adapted Core Prescribing Indicators (N = 151)

| Indicator | Value |
| --- | --- |
| Average number of drugs prescribed per patient (all categories) | 10.56 ± 2.19 |
| Average number of anticancer drugs per patient | 1.72 ± 0.67 |
| Patients prescribed combination anticancer therapy | 60.3% (91/151) |
| Patients prescribed anticancer monotherapy | 39.7% (60/151) |
| Patients with an antibiotic co-prescribed | 2.0% (3/151) |
| Patients with an additional injection prescribed | 6.6% (10/151) |
| Patients with a cytoprotective drug prescribed | 27.2% (41/151) |

*Note. Values are M ± SD or % (n/N). Indicators are adapted from, but not identical to, the standard WHO/INRUD core prescribing indicator set; anticancer-specific and regimen-complexity measures replace the standard % generic and % essential-medicines-list indicators, which are reported separately in Table 3.*

#### Prescribing Indicators

On average, patients received 1.72 anticancer drugs (SD = 0.67, range 1–4), of which 1.60 (SD = 0.68) were administered intravenously and 0.11 (SD = 0.32) orally. Patients additionally received an average of 3.70 injectable adjuvant drugs (SD = 1.15, range 0–7) and 5.15 oral supportive drugs (SD = 1.45, range 0–8), yielding a mean of 10.56 total drugs prescribed per patient (SD = 2.19, range 3–16). Prescribing quality indicators showed a very high rate of generic prescribing, with an average of 99.85% of drugs per patient prescribed by generic name (SD = 1.08%), and 98.0% of patients (n = 148) having all drugs prescribed generically. Adherence to the Nepal National List of Essential Medicines (NLEM) 2021 was more variable: on a strict matching criterion, an average of 63.17% of drugs per patient appeared on the NLEM (SD = 20.28%), and only 3.3% of patients (n = 5) had every drug on the strict NLEM list. When borderline matches were included, the average rose to 67.67% of drugs per patient (SD = 17.56%). Antibiotic co-prescription was rare, occurring in only 2.0% of patients (n = 3), and should be interpreted with caution given the very small cell size. An additional (non-chemotherapy) injection was used in 6.6% of patients (n = 10), and a cytoprotective drug was co-prescribed in 27.2% of patients (n = 41).

### Cost Analysis of Anticancer Therapy

#### Cost Variables

The average cost of anticancer therapy was NPR 15,245.28 per patient, ranging from NPR 1,665.60 to NPR 101,251.96. The average maximum retail price was NPR 38,803.22, and the average Nepal Health Insurance Board reimbursement rate was NPR 25,852.36. Cost varied widely across patients, with a small number of high-cost patients raising the average well above the typical (median) cost of NPR 11,196.00 (Table 5). A log-transformed cost variable (LogCost) was used for the comparisons and regression analysis below, since it more closely approximated a normal distribution than the raw cost values (Additional File 1, Table S3).

**Table 5.** Cost of Anticancer Drug Therapy per Patient (NPR)

| Cost measure | Mean ± SD | 95% CI | Median | Min–Max |
| --- | --- | --- | --- | --- |
| Total actual cost | 15,245.28 ±<br>15,672.33 | 12,725.22,<br>17,765.35 | 11,196.00 | 1,665.60–<br>101,251.96 |
| Total maximum<br>retail price (MRP) | 38,803.22 ±<br>29,157.04 | 34,114.85,<br>43,491.59 | 32,556.48 | 5,426.08–<br>200,066.93 |
| Total cost, Health<br>Insurance Board<br>rate | 25,852.36 ±<br>21,778.96 | 22,350.36,<br>29,354.36 | 21,156.18 | 3,526.03–<br>145,886.82 |

Note. Mean, SD, median, minimum, and maximum are from the Section 3 DESCRIPTIVES output. The 95% CI for Total\_Cost is taken directly from the EXAMINE procedure; the 95% CIs for MRP and the HIB rate were calculated as $\text{Mean} \pm t_{0.975, 150} \times (SD/\sqrt{N})$ , using the critical value ( $t \approx 1.976$ , $df = 150$ ) implied by the EXAMINE-derived CI for Total\_Cost. IQR was not printed in the available output and is therefore omitted rather than estimated.

#### Bivariate Associations with Cost

Cost was not related to patient age, chemotherapy cycle number, the number of drugs prescribed, or the percentage of drugs that were generic or on the essential medicines list. However, cost was higher among male patients than female patients and among patients whose drugs were not fully on the essential medicines list; it did not differ by antibiotic co-prescription, additional injection use, cytoprotective drug use, regimen type, or generic prescribing status (Additional File 1, Tables S1–S2).

#### Multiple Linear Regression Predicting Log-Transformed Cost

A multiple linear regression examined the combined contribution of patient age, sex, chemotherapy cycle number, regimen complexity, total anticancer drugs per patient, total drugs prescribed per patient, antibiotic co-prescription, additional injection use, cytoprotective drug use, percentage of drugs on the strict NLEM list, and organ system to cost. Together, these variables accounted for about 11% of the variation in cost (Table 6). None of the individual variables was independently associated with cost (Table 6).

**Table 6.** Summary of Multiple Regression Coefficients Predicting LogCost (N = 151)

| Predictor | $\beta$ | t | p | 95% CI |
| --- | --- | --- | --- | --- |
| (Constant) | — | 13.25 | <.001 | [7.591, 10.257] |
| Patient age | .107 | 1.15 | .254 | [-0.004, 0.016] |
| Sex (Female = 1) | -.068 | -0.61 | .543 | [-0.466, 0.246] |
| Chemotherapy cycle number | .076 | 0.89 | .375 | [-0.033, 0.085] |
| Regimen: Combination = 1 | -.065 | -0.31 | .755 | [-0.729, 0.529] |
| Total anticancer drugs / patient | -.070 | -0.31 | .755 | [-0.574, 0.418] |
| Total drugs prescribed / patient | .086 | 0.89 | .378 | [-0.037, 0.097] |
| Antibiotic co-prescribed | -.159 | -1.50 | .135 | [-1.981, 0.269] |
| Additional injection used | -.124 | -1.43 | .155 | [-0.889, 0.143] |
| Cytoprotective drug used | .066 | 0.64 | .521 | [-0.229, 0.451] |
| % drugs on NLEM, strict | -.054 | -0.60 | .553 | [-0.008, 0.004] |
| Organ system:<br>Gastrointestinal | .029 | 0.24 | .812 | [-0.425, 0.541] |
| Organ system:<br>Gynecological | -.036 | -0.30 | .768 | [-0.491, 0.363] |
| Organ system: Head & Neck | .026 | 0.19 | .849 | [-0.548, 0.666] |
| Organ system: Respiratory | .091 | 0.72 | .472 | [-0.320, 0.688] |
| Organ system: Others<br>(Genitourinary/Hematologic) | .093 | 0.80 | .425 | [-0.432, 1.020] |

Note. Reference categories: Sex = Male; Regimen = Monotherapy; Organ system = Breast. Dependent variable:
LogCost. Overall model: $R^2 = .113$ , adjusted $R^2 = .014$ , $F(15,135) = 1.15$ , $p = .323$ . VIF = variance inflation factor.
95% CI for B calculated as $B \pm t_{0.975,135} \times SE B$ ( $t_{0.975,135} = 1.978$ ).

## Discussion

Among 151 chemotherapy patients at this tertiary cancer hospital, prescribing was characterized by a modest number of anticancer agents per patient (mean 1.72), a substantially larger burden of adjuvant and supportive drugs, near-universal generic prescribing (99.85%), and only partial adherence to the national essential medicines list (63–68%). Mean actual chemotherapy cost per patient (NPR 15,245) was well below mean maximum retail price (NPR 38,803), and cost was highly right-skewed. In bivariate analysis, LogCost was higher among male patients and among patients not fully NLEM-compliant, but no patient, disease, or prescribing variable — including sex — independently predicted LogCost in an overall regression model that did not reach statistical significance.

The demographic and diagnostic profile observed here differs from other Nepalese anticancer drug utilization studies largely as a function of case mix. Our sample was older on average and markedly female-predominant (67.5%), consistent with the predominance of breast and gynecological malignancies (49.0% combined); by contrast, a lung-cancer-focused drug utilization study at the same hospital reported a younger, male-predominant sample (mean age 58.21 ± 13.22 years, 57% male)[6], and an adverse-drug-reaction study across a broader Nepalese oncology sample reported a lower mean age (49.93 ± 14.27 years)[5]. This suggests that patient- and cost-related findings in anticancer drug utilization studies in Nepal may be strongly shaped by the diagnostic mix of the sampled hospital or ward, and comparisons across studies should account for this.

The prescribing volume observed here (10.56 total drugs and 1.72 anticancer drugs per patient) sits within the range reported by other hospital-based anticancer drug utilization studies in South Asia, which have found 8.77–9.28 total drugs per encounter at other Nepalese and Indian centers [6,12]and up to 12.22 total drugs with 1.73 anticancer drugs per encounter in an Indian government hospital treating metastatic disease[12], as well as 1.72–2.11 anticancer drugs per encounter in tertiary Indian oncology settings [26]. Beyond prescribing volume itself, drug wastage has been identified as a related cost driver in comparable settings, with one Indian tertiary-hospital study quantifying substantial financial loss from chemotherapy drug wastage in paediatric cancer care[27]. This convergence across independently conducted studies, despite differing case mixes and health systems, suggests that a total prescribing burden of roughly 9–13 drugs per chemotherapy encounter — combining cytotoxic agents with antiemetics, gastroprotective drugs, and other supportive therapy — may be a broadly representative feature of real-world combination chemotherapy in South Asian tertiary care, rather than an artifact specific to this hospital.

By contrast, the WHO/INRUD literature outside oncology — spanning general outpatient and primary-care settings in Ethiopia, Pakistan, and Nigeria — typically reports considerably lower average drugs per encounter (1.75–3.4) alongside more variable antibiotic and injection use [7–9]. The higher drug burden in the present sample is therefore best interpreted as a feature of oncology prescribing specifically, where antiemetic, gastroprotective, and other supportive medications are added systematically alongside the anticancer regimen itself, rather than evidence of generally excessive prescribing; this is consistent with adverse-drug-reaction surveillance data from Eastern Indian cancer centers showing that supportive medications are integral to managing the substantial toxicity burden of cytotoxic chemotherapy[15,16].

Generic prescribing in this sample was essentially complete (99.85% of drugs, 98.0% of patients), a favorable prescribing-quality indicator consistent with WHO/INRUD expectations for rational drug use[3]. Adherence to the NLEM 2021 was more variable, with only 3.3% of patients fully compliant under a strict matching criterion. Because the current NLEM explicitly includes a section on antineoplastic and immunomodulating agents [4], this gap may partly reflect the narrower scope of that list relative to the range of regimens required for the diverse cancer types treated at this hospital, rather than irrational prescribing per se; it nonetheless represents an actionable target for future formulary and procurement review.

This pattern is broadly consistent with international observations that essential-medicines-list adherence for cancer medicines lags behind adherence for medicines used to treat other conditions: a survey of oncologists across 82 countries found inconsistent real-world availability even for medicines respondents themselves regarded as essential[22], and a comparative analysis found that national essential medicines lists in many low- and middle-income countries capture well under half of guideline-recommended oncology medicines[23]. The WHO’s own selection criteria for cancer medicines have become progressively more stringent over successive Model List revisions[21], which may itself widen, rather than narrow, the gap between what guidelines recommend, what a national list includes, and what a given hospital actually prescribes — precisely the gap this study’s NLEM-adherence indicators are intended to surface locally.

The gap between mean actual cost (NPR 15,245) and mean maximum retail price (NPR 38,803) observed here is consistent with documented brand-level price variation in the Nepalese anticancer medicine market, where price differences between brands of the same molecule have been reported to exceed 300%[20], and with the partial buffering role of the Health Insurance Board reimbursement mechanism (mean NPR 25,852 in this sample). These patient-level costs are considerably smaller in magnitude than the whole-course direct cost of cancer treatment reported elsewhere in Nepal (mean NPR 387,500 per patient, of which medical costs comprised over 80%) [19], reflecting the difference between a single chemotherapy encounter and a full multi-modality treatment course; taken together, however, both findings point to substantial and variable out-of-pocket exposure for Nepalese cancer patients.

These patient-level cost findings echo the broader financial-toxicity literature: landmark work defining and measuring financial toxicity in insured cancer patients in high-income settings found that a majority reported meaningful financial burden despite insurance coverage[28–30], and out-of-pocket cost reviews spanning multiple income settings consistently find that medication costs, rather than consultation or diagnostic fees, dominate the direct cost of cancer care[31] — a pattern directly mirrored here, where anticancer drug cost, rather than any other component of care, was the object of measurement. In the largely uninsured, out-of-pocket-dominated Nepalese context, where the Health Insurance Board mechanism observed in this sample offsets only part of the gap between actual and listed prices, the combination of high MRP-to-paid-cost variability[20] and limited catastrophic-expenditure protection[19] plausibly compounds financial toxicity risk beyond what is described in insured, high-income-country cohorts.

This pattern mirrors a much larger international literature on the financial toxicity of cancer care: out-of-pocket costs have been shown to impose a substantial and often catastrophic burden on patients across income settings, from the United States and Canada to Western Europe and Australia[32], with landmark studies coining and operationalizing the term “financial toxicity” to describe how treatment-related costs can rival clinical toxicity as a source of patient distress and even treatment non-adherence[30]. A systematic review and meta-analysis focused specifically on low- and middle-income countries found pooled prevalence estimates of objective financial toxicity ranging from roughly 18% to over 90% depending on cancer type and setting [33], and a Malaysian study of urologic cancer patients similarly found that both objective and subjective financial toxicity were associated with poorer quality of life[34] — findings broadly consistent with the high mean chemotherapy cost relative to typical household income observed in the Nepalese context[19].

The attenuation of the sex difference in LogCost between the unadjusted (bivariate) and adjusted (regression) analyses, together with the non-significant overall regression model, suggests that the patient- and prescribing-level variables captured in this study do not by themselves explain much of the variation in chemotherapy cost. This is plausible given that anticancer drug pricing in Nepal varies considerably by brand and manufacturer rather than by patient characteristics[20]; regimen- and drug-selection-level cost drivers not captured here (for example, brand choice, originator versus generic formulation, and negotiated institutional pricing) may be more informative predictors of cost than the demographic and prescribing variables examined in this model.

### Strengths and limitations of the study

This study jointly characterized anticancer drug utilization and cost at the level of the individual chemotherapy patient across a broad mix of cancer diagnoses, an uncommon combination in the existing Nepal-specific literature, which has more often focused on either a single tumor type or on adverse drug reactions rather than cost. Several limitations should be considered. The study was single-center and cross-sectional, precluding causal inference and limiting generalizability to other Nepalese cancer hospitals. The achieved sample (151 patients over a two-month window) corresponded to a response rate of approximately 95.6% relative to the non-response-adjusted minimum target of 158 patients; however, the planned one-way ANOVAs across diagnosis, organ system, age band, ethnicity, and cycle-count group could not be completed in the current analysis and are being re-run. Several patient-level indicators had very small numbers of positive cases (antibiotic co-prescription, n = 3; strict NLEM compliance, n = 5), and comparisons involving these indicators should be interpreted cautiously. Cost data were restricted to the recorded actual cost, maximum retail price, and Health Insurance Board rate for anticancer drugs and did not capture indirect or non-medical costs (for example, travel, lodging, or lost income), which other Nepalese cost-of-cancer studies suggest can be substantial. Finally, a planned non-parametric comparison of untransformed cost by sex, regimen type, and cytoprotective-drug use was not completed in the current analysis and is a further target for revision.

Consistent with methodological guidance from systematic reviews of cancer cost-of-illness studies more broadly, the cost measures used here (actual cost, MRP, and HIB rate) capture direct medical drug cost only; they do not capture indirect costs such as travel, lost wages, or informal caregiving time, which other financial-toxicity literature identifies as substantial additional contributors to the total economic burden of cancer care.

### Study implications

These findings support the continued use of WHO/INRUD-style prescribing indicators as a routine monitoring tool in oncology settings, given their ability to flag both prescribing strengths (near-universal generic use) and gaps (partial essential-medicines-list adherence) that might otherwise go unquantified. They also support institutional and policy attention to anticancer drug pricing and procurement — particularly the gap between listed and paid prices — as a lever for reducing the financial burden of chemotherapy on patients, independent of any single patient or prescribing characteristic.

More broadly, the combination of near-complete generic prescribing with only partial essential-medicines-list adherence observed here is a pattern also seen internationally as national lists struggle to keep pace with evolving guideline recommendations; periodic realignment of hospital procurement and the national list — informed by structured audits of the kind conducted here — remains one of the most consistently recommended, low-cost levers for improving rational medicine use in resource-limited settings.

## Conclusion

In this descriptive cross-sectional study of 151 chemotherapy patients at a tertiary cancer hospital in Nepal, anticancer drug prescribing was characterized by near-universal generic use but only partial adherence to the national essential medicines list, and the cost of anticancer drug therapy varied widely and was not reliably explained by the patient, disease, or prescribing factors examined. Continued patient-level monitoring of prescribing pattern and cost, alongside efforts to align anticancer procurement more closely with the essential medicines list and to address brand-level price variation, may help contain the financial burden of chemotherapy in this setting.

## Data Availability

The minimal dataset underlying the findings of this study will provided by the crossponding author.

## Acknowledgments

We would like to express our sincere gratitude to the Masters of Pharmaceutical Sciences Program, CiST College, Pokhara University, for providing the opportunity to complete this study as partial fulfillment of the requirements for the Master degree in Clinical Pharmacy.

We are deeply grateful to our supervisor, Ms. Poonam Pant, co-supervisor, Dr. Prayas Ghimire, Prof. Naveen Shrestha, for their supervision, guidance, valuable support, constructive ideas, and feedback throughout this research.

We would like to thank Prof. Dr. Sudhamsu KC, Dr. Dilip Sharma, and all the staff of the Centre for Liver Disease, Kathmandu, for their approval, guidance, support, and for providing the patient-related information required to complete our thesis.

We are also thankful to all the patients and their visiting parties for their kind and supportive behavior.

Finally, we would like to express our gratitude to everyone who directly or indirectly provided their time, support, and guidance for the successful completion of our thesis

## Supporting information

S1 Table. Pearson correlations between LogCost and continuous predictors (N = 151).

S2 Table. Independent-samples t tests for LogCost by binary indicators (N = 151).

S3 Table. Normality testing of continuous variables (as reported in the output).

Additional file 1 — for separate Supporting Information upload

## Declarations

### Ethics approval and consent to participate

Ethical approval was obtained from the Institutional Review Committee of CIST College, Pokhara University (Approval No. IRC/114/078/79), with administrative approval from Bhaktapur Cancer Hospital. Written and verbal informed consent were obtained from participants where applicable; participation was voluntary and confidentiality was maintained throughout.

### Consent for publication

Not applicable — no individual patient data, images, or identifying information are presented.

### Availability of data and materials

The dataset analyzed in this study is available from the corresponding author on reasonable request.

